# Contamination of air and surfaces in workplaces with SARS-CoV-2 virus: a systematic review

**DOI:** 10.1101/2021.01.25.21250233

**Authors:** JW Cherrie, MPC Cherrie, A Davis, D Holmes, S Semple, S Steinle, E MacDonald, G Moore, M Loh

**Affiliations:** Institute of Occupational Medicine, Edinburgh EH14 4AP, UK; Heriot Watt University, Institute of Biological Chemistry Biophysics and Bioengineering Edinburgh EH14 4AS, UK; University of Edinburgh, School of Geosciences, Drummond St, Edinburgh EH8 9XP, UK; University of Stirling, Institute for Social Marketing and Health, Stirling FK9 4LA, UK; University of Glasgow, Institute of Health and Wellbeing, Glasgow UK; National Infection Service, Public Health England, Porton Down, Salisbury SP4 0JG, UK

**Keywords:** SARS-CoV-2, virus, aerosol, surface, fomite, hospital, transportation

## Abstract

**Objectives:** This systematic review aimed to evaluate the evidence for air and surface contamination of workplace environments with SARS-CoV-2 RNA and the quality of the methods used to identify actions necessary to improve the quality of the data.

**Methods:** We searched Web of Science and Google Scholar until 24^th^ December 2020 for relevant articles and extracted data on methodology and results.

**Results:** The vast majority of data come from healthcare settings, with typically around 6 % of samples having detectable concentrations of SARS-CoV-2 RNA and almost none of the samples collected had viable virus. There were a wide variety of methods used to measure airborne virus, although surface sampling was generally undertaken using nylon flocked swabs. Overall, the quality of the measurements was poor. Only a small number of studies reported the airborne concentration of SARS-CoV-2 virus RNA, mostly just reporting the detectable concentration values without reference to the detection limit. Imputing the geometric mean air concentration assuming the limit of detection was the lowest reported value, suggests typical concentrations in health care settings may be around 0.01 SARS-CoV-2 virus RNA copies/m^3^. Data on surface virus loading per unit area were mostly unavailable.

**Conclusion:** The reliability of the reported data is uncertain. The methods used for measuring SARS-CoV-2 and other respiratory viruses in work environments should be standardised to facilitate more consistent interpretation of contamination and to help reliably estimate worker exposure.

**Key messages:**

1. What is already known about this subject?
  - Low level contamination of air and surfaces in hospitals with SARS-CoV-2 RNA have been reported during the Covid-19 pandemic.
  - Limited data have published from non-healthcare settings.
2. What are the new findings?
  - Typically, around 6% of air and surface samples in hospitals were positive for SARS-COV-2 RNA, although there is very limited data for non-healthcare settings.
  - The quality of the available measurement studies is generally poor, with little consistency in the sampling and analytical methods used.
  - Few studies report the concentration of SARS-CoV-2 in air or as surface loading of virus RNA, and very few studies have reported culture of the virus.
  - The best estimate of typical air concentrations in health care settings is around 0.01 SARS-CoV-2 virus RNA copies/m^3^
3. How might this impact on policy or clinical practice in the foreseeable future?
  - There should be concerted efforts to standardise the methods used for measuring SARS-CoV-2 and other respiratory viruses in work environments.

## INTRODUCTION

A large-scale, global research effort has been directed at understanding the risks from Covid-19 infections and seeking successful clinical interventions to help patients. There have been almost 70,000 scientific papers published on the topic during the first 10 months of 2020, around 2.3% of all scientific publications during this period ^a^ . Despite all this new knowledge there has been little quantitative data on the extent of exposure to SARS-CoV-2 of workers in the healthcare sector, and much debate about the best strategies to protect them from infection^1 2^. The relative contribution of different transmission routes in terms of the risk of workplace infection continues to be poorly understood^3^.

Viruses may be transmitted from an infected patient to healthcare workers through a number of routes: by large droplets emitted from coughs or sneezes that may splatter directly on the worker’s face; from fomite transmission where the worker contacts a surface contaminated by droplet emission and then transfers virus from the surface to their nose, mouth or eyes; and finally, from aerosol transmission where fine particles containing the virus are emitted from the respiratory system of the patient, become airborne for a period and may then be inhaled by the worker. The relative importance of these three routes in determining the risk of infection is poorly understood for SARS-CoV-2^3^.

In the early stages of the pandemic, the World Health Organisation (WHO) was clear that “SARS-CoV-2 transmission appears to mainly be spread via droplets and close contact with infected symptomatic cases” and in most circumstances aerosol transmission was considered unlikely^4^. However, as knowledge of the virus has increased it has become apparent that aerosol transmission may be more important than was previously thought and some have argued that it is a major source of infection^5^.

The situation is further complicated because our understanding of the extent of SARS-CoV-2 air and surface contamination in hospitals and other workplaces is limited. There are only around 0.06% of all the Covid-19 related research papers that describe measurements of environmental contamination^b^, and these data tend not to have been appropriately summarised. Without an evidence base to understand how exposure or transmission takes place it is difficult to set out rational plans to control SARS-CoV-2 in the workplace. For example, this has resulted in heated policy debates about whether it is necessary to wear effective respiratory protection when there are no deliberate aerosol generating procedures on Covid-19 patients. It is also likely that the relative importance of different transmission routes will vary depending on the workplace, the tasks being performed and the interaction with an infective source.

The aim of this review is to summarise the reported SARS-CoV-2 RNA air and surface contamination concentrations in workplace settings where the virus is present, particularly considering the quality of the methods used, to draw lessons for future methodological developments.

## METHODS

We searched Web of Science (WoS) using the terms in the title ((SARS-CoV-2 or “severe acute respiratory syndrome”) and air), and ((SARS-CoV-2 or “severe acute respiratory syndrome”) and surface), for all languages and all document types. In addition, we searched the Google Scholar database for the above search terms, excluding the phrase “severe acute respiratory syndrome” to restrict the hits to a manageable number. The references were combined into a single database and duplicate entries were removed. The entries were then screened by a single researcher (Cherrie, JW) on the basis of title and abstract to identify informative papers containing data on either air or surface concentrations of SARS-CoV-2 RNA in workplaces, including papers that reported their results as either positive or negative contamination without quantifying the extent of the contamination; papers not in the English language were excluded. Data on other measures, for example virus RNA in exhaled breath condensate, were excluded. Following the initial literature search we set up a Google Scholar alert using the same search terms as used initially. These produced periodic updates that were screened in the same way as the original citations and relevant publications were added to the final list. These periodic updates were included up to the 24^th^ December 2020. Copies of all papers were obtained, and data extracted into tables for summarisation. Numeric data extraction was checked by a second researcher (Steinle).

Data were summarised graphically using the DataGraph software. For datasets with more than one detectable result in a dataset of 10 or more measurements we used the elnormCensored function in in the R-package EnvStats v2.3.1 to estimate the geometric mean and associated 95% confidence intervals using the maximum likelihood method.

## RESULTS

The initial WoS searches identified 44 papers relating to airborne contamination and 42 on surface contamination, some of which were included in both lists. Google Scholar produced a greater number of references: 137 on air contamination and 80 relating to surface contamination (Figure 1).

**Figure 1:**
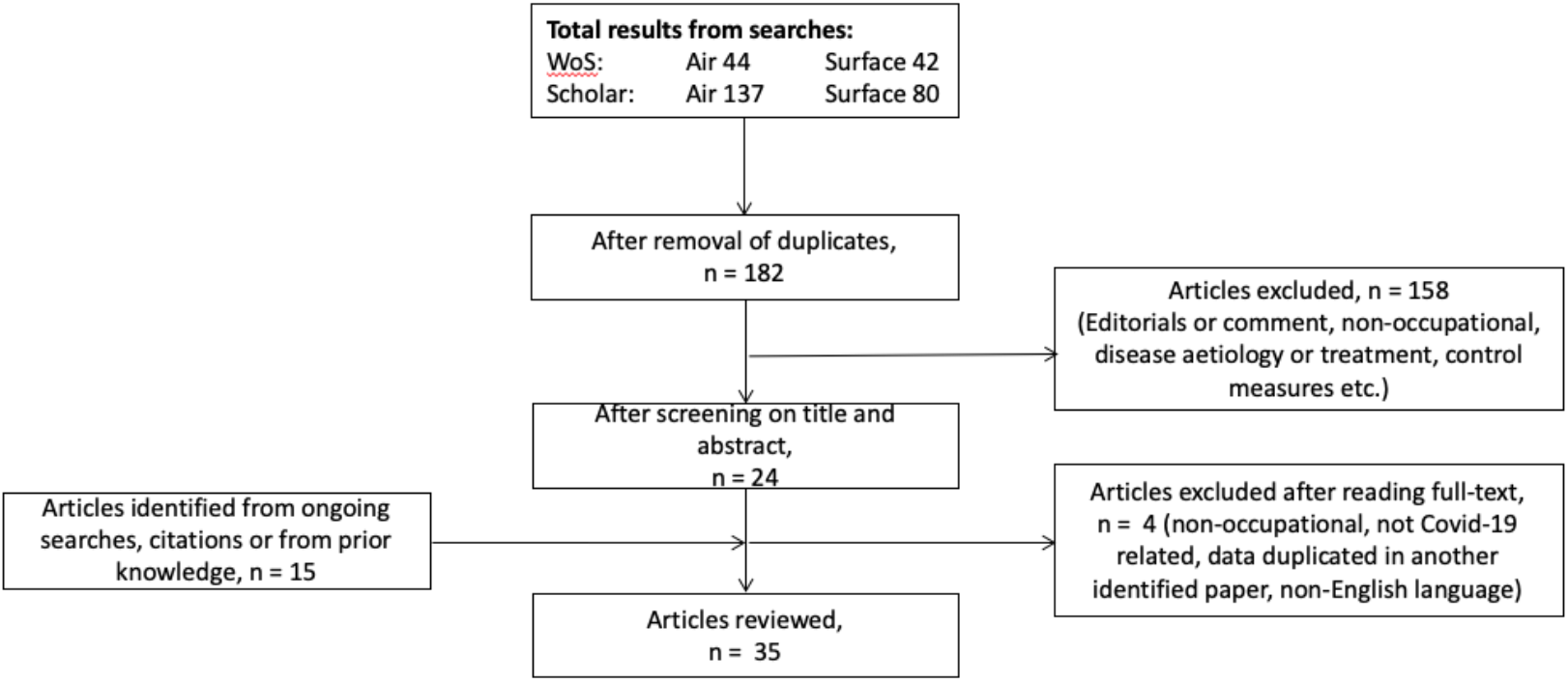
Results from the systematic literature search.

After the removal of duplicates there were 182, which resulted in 26 informative papers for inclusion in the review. A further thirteen papers were added from the ongoing literature searches or other sources and on further reading four papers were excluded: one duplicated data in another identified paper, one related to non-occupational exposure, one was not related to Covid-19 infection risks and the last was written in Persian. In the end, 35 papers were reviewed: three were available as pre-prints and the remainder as peer-reviewed publications (Table 1).

**Table 1:**
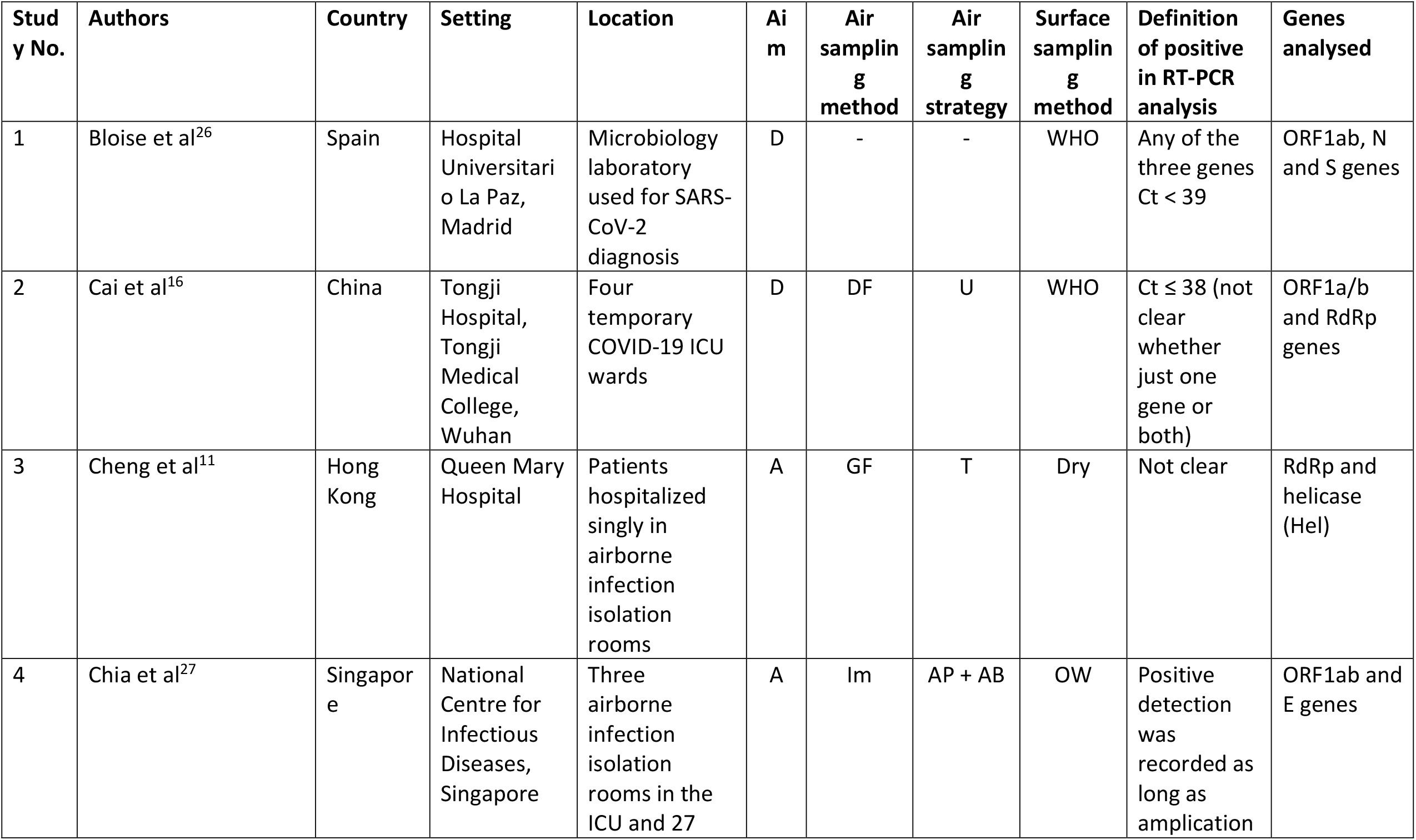
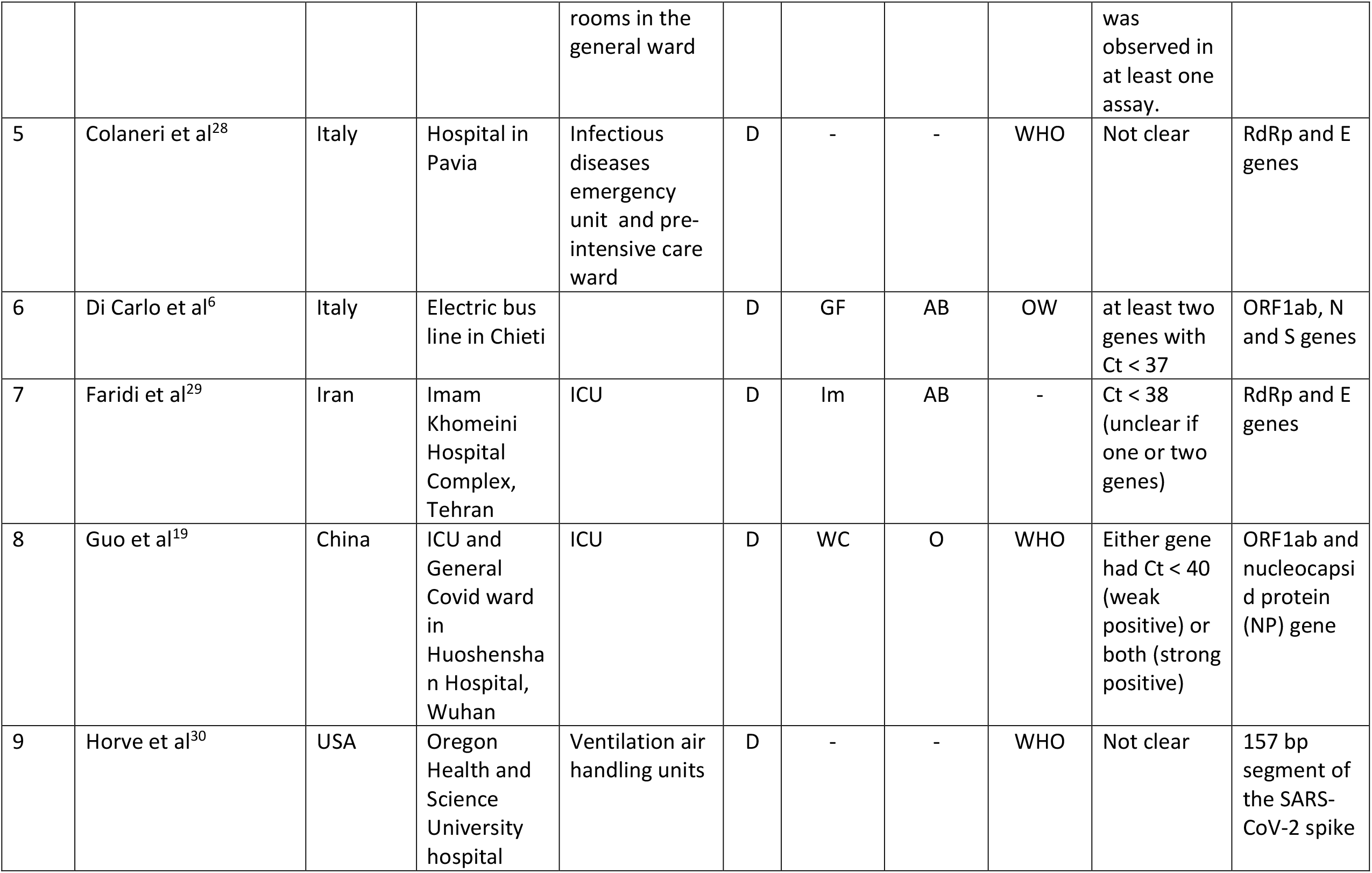
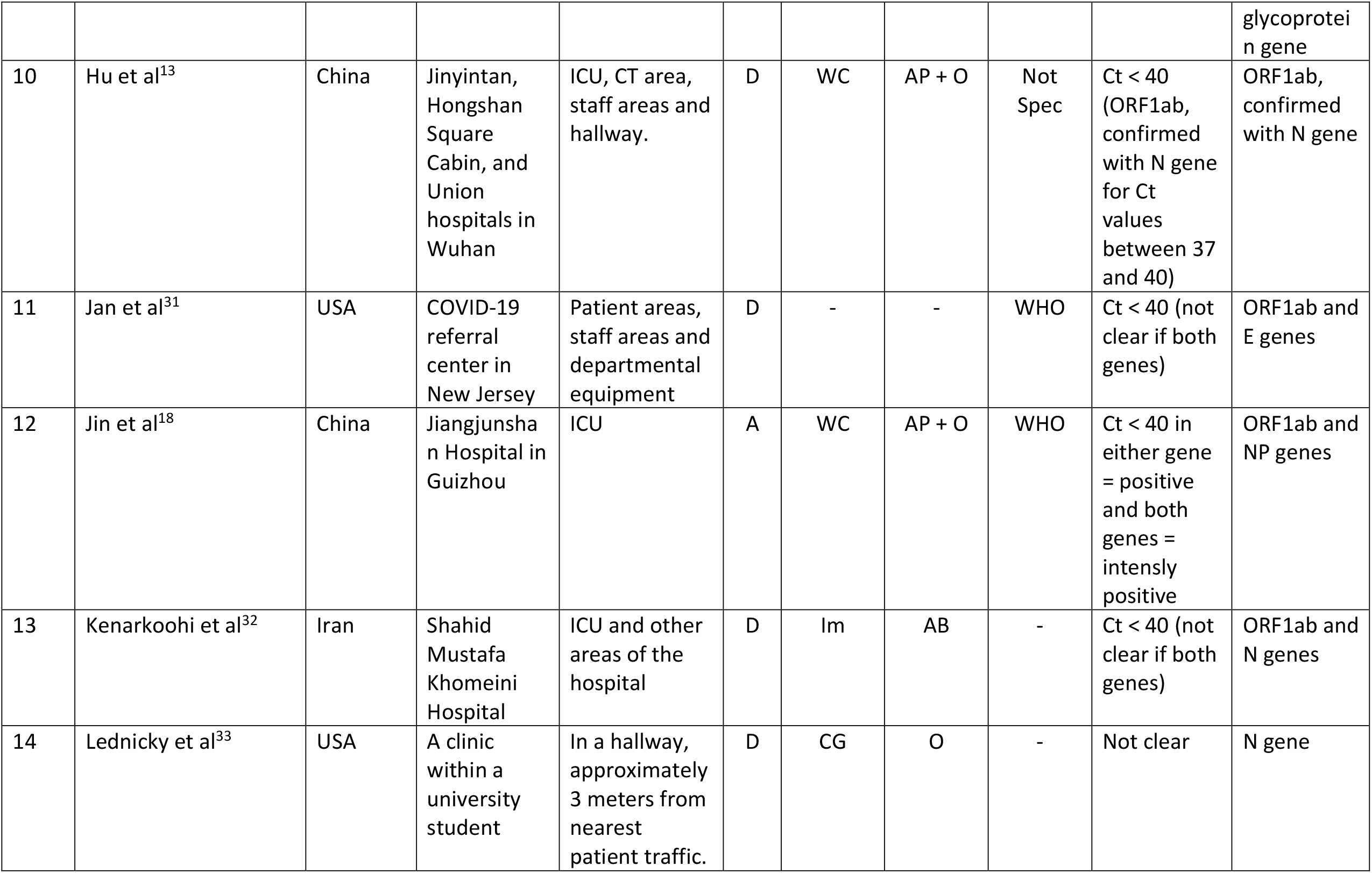
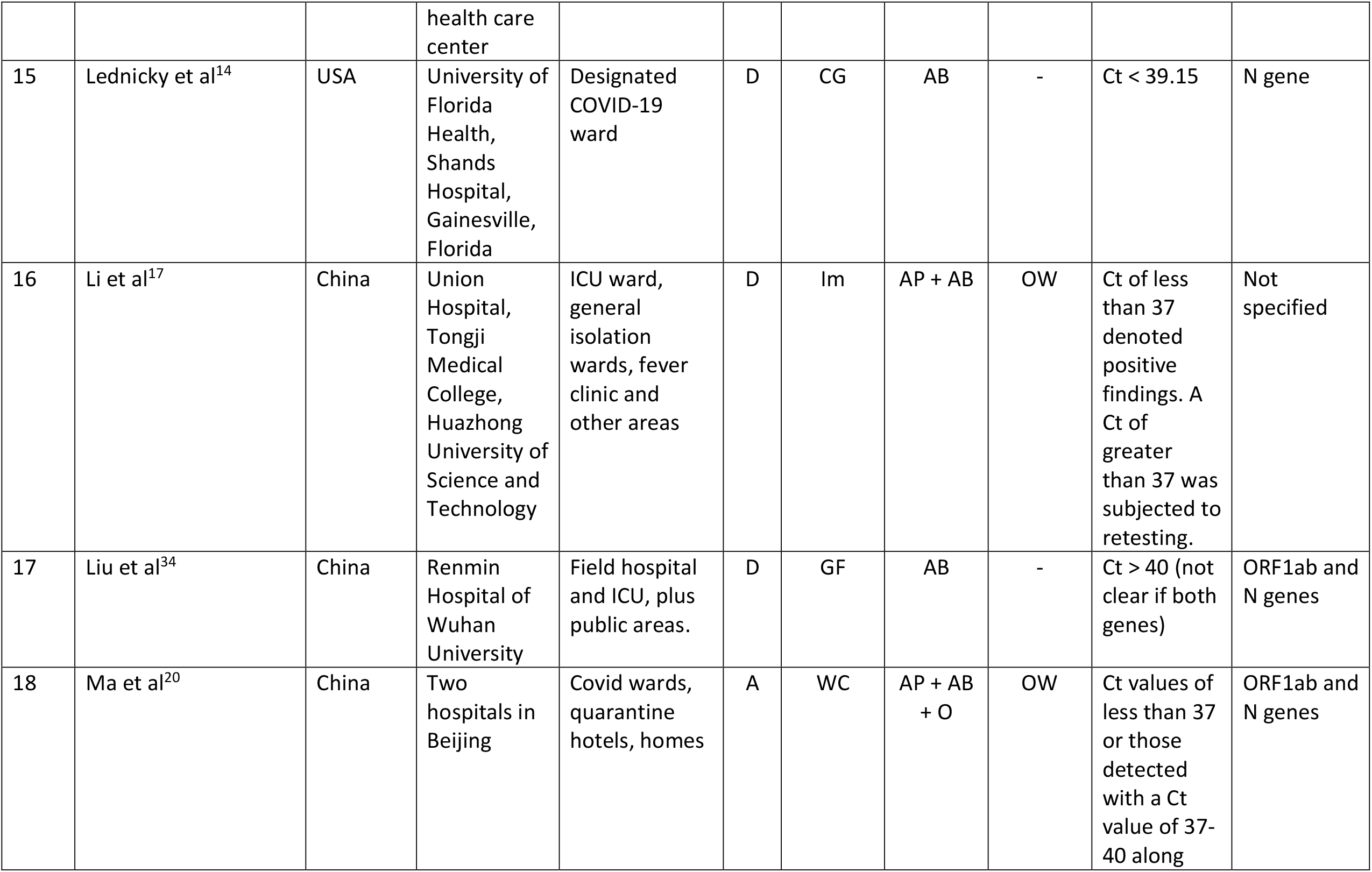
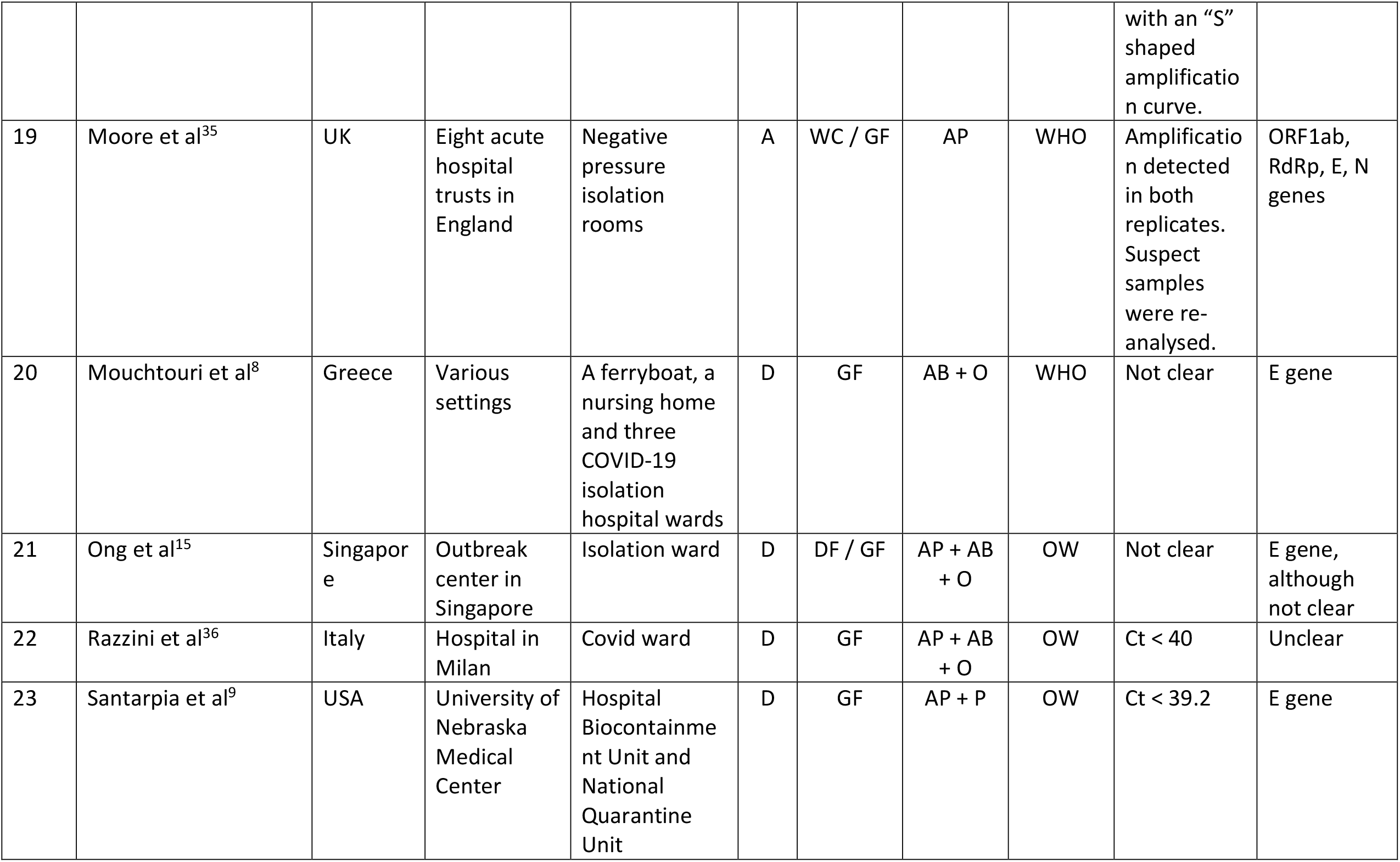
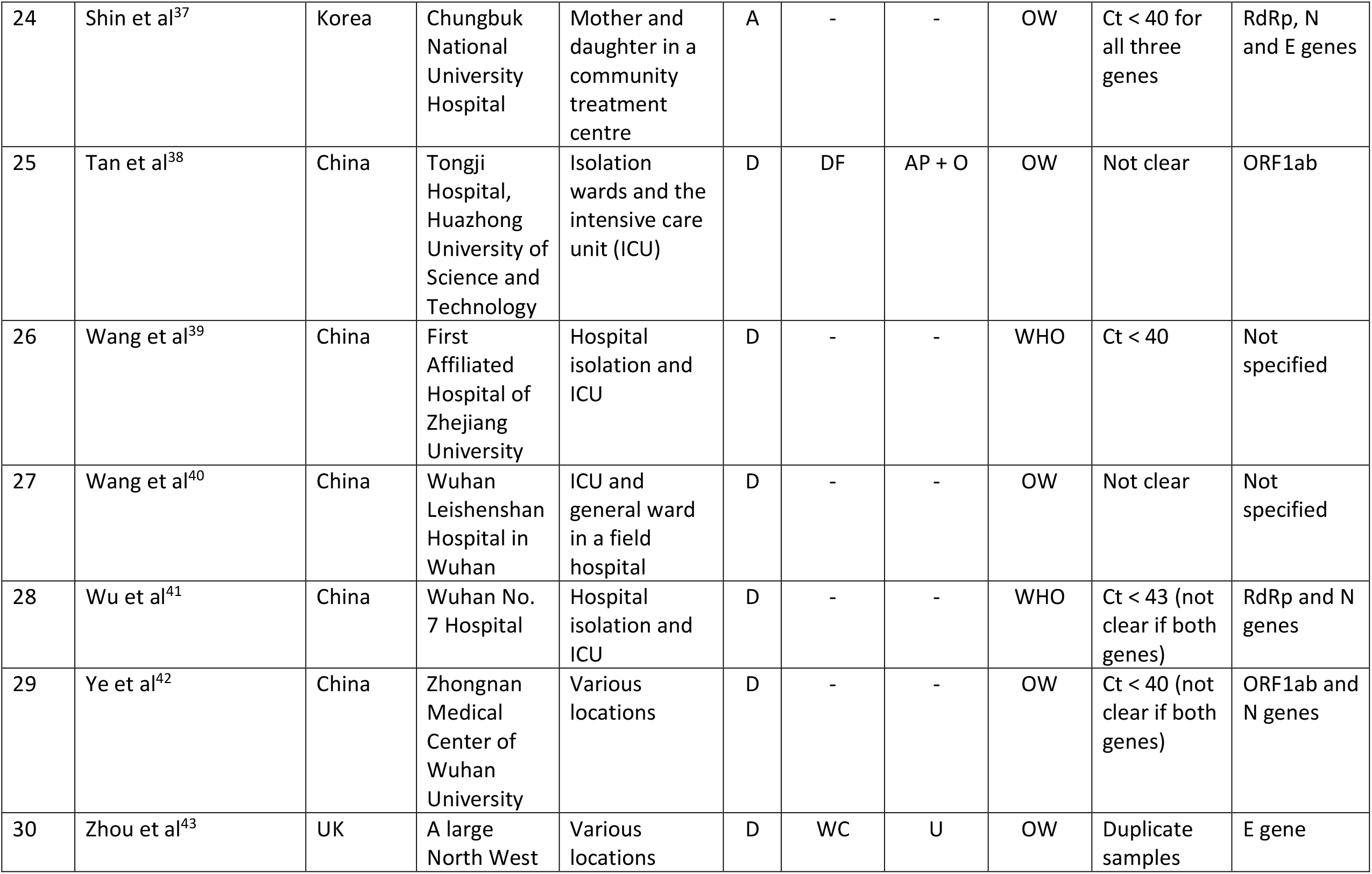
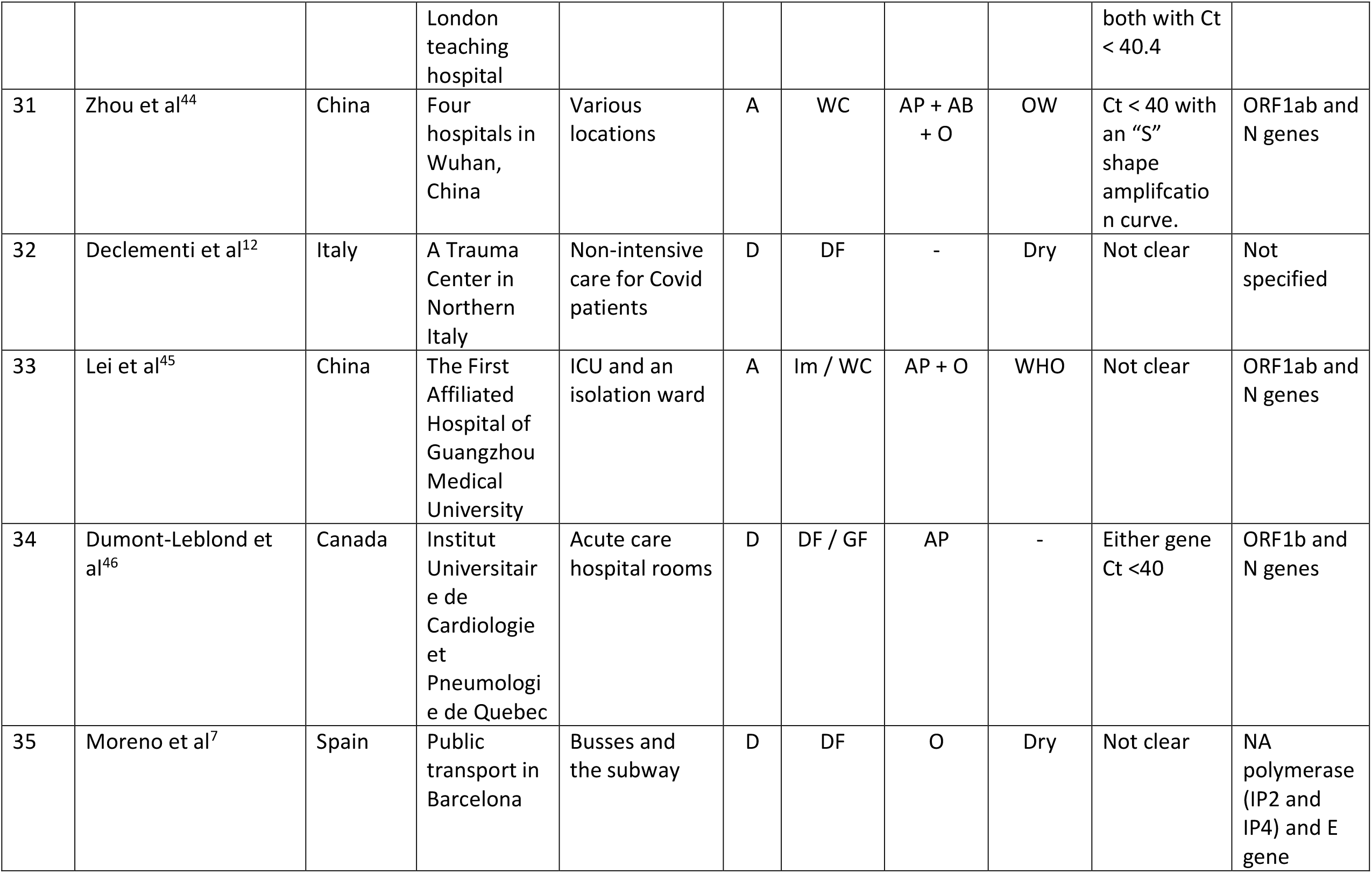
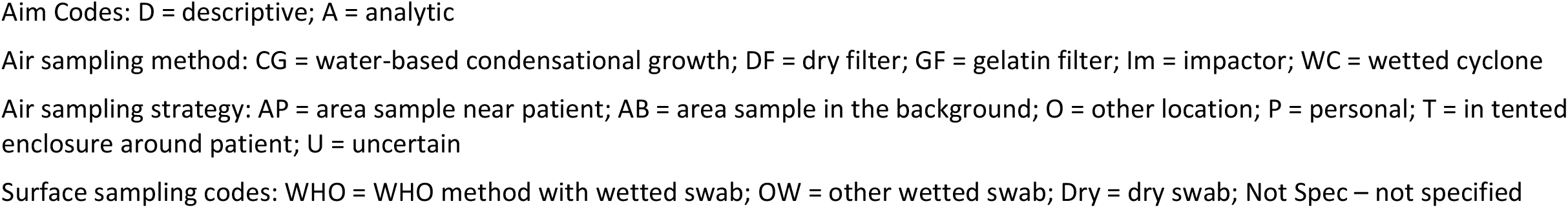
Summary of studies reviewed: setting, study aims and methodology.

| Study No. | Authors | Country | Setting | Location | Aim | Air sampling method | Air sampling strategy | Surface sampling method | Definition of positive in RT-PCR analysis | Genes analysed |
| --- | --- | --- | --- | --- | --- | --- | --- | --- | --- | --- |
| 1 | Bloise et al<sup>26</sup> | Spain | Hospital Universitario La Paz, Madrid | Microbiology laboratory used for SARS-CoV-2 diagnosis | D | - | - | WHO | Any of the three genes<br>Ct < 39 | ORF1ab, N and S genes |
| 2 | Cai et al<sup>16</sup> | China | Tongji Hospital, Tongji Medical College, Wuhan | Four temporary COVID-19 ICU wards | D | DF | U | WHO | Ct ≤ 38 (not clear whether just one gene or both) | ORF1a/b and RdRp genes |
| 3 | Cheng et al<sup>11</sup> | Hong Kong | Queen Mary Hospital | Patients hospitalized singly in airborne infection isolation rooms | A | GF | T | Dry | Not clear | RdRp and helicase (Hel) |
| 4 | Chia et al<sup>27</sup> | Singapore | National Centre for Infectious Diseases, Singapore | Three airborne infection isolation rooms in the ICU and 27 | A | Im | AP + AB | OW | Positive detection was recorded as long as amplification | ORF1ab and E genes |
|  |  |  |  | rooms in the general ward |  |  |  |  | was observed in at least one assay. |  |
| 5 | Colaneri et al<sup>28</sup> | Italy | Hospital in Pavia | Infectious diseases emergency unit and pre-intensive care ward | D | - | - | WHO | Not clear | RdRp and E genes |
| 6 | Di Carlo et al<sup>6</sup> | Italy | Electric bus line in Chieti |  | D | GF | AB | OW | at least two genes with Ct < 37 | ORF1ab, N and S genes |
| 7 | Faridi et al<sup>29</sup> | Iran | Imam Khomeini Hospital Complex, Tehran | ICU | D | Im | AB | - | Ct < 38 (unclear if one or two genes) | RdRp and E genes |
| 8 | Guo et al<sup>19</sup> | China | ICU and General Covid ward in Huoshenshan Hospital, Wuhan | ICU | D | WC | O | WHO | Either gene had Ct < 40 (weak positive) or both (strong positive) | ORF1ab and nucleocapsid protein (NP) gene |
| 9 | Horve et al<sup>30</sup> | USA | Oregon Health and Science University hospital | Ventilation air handling units | D | - | - | WHO | Not clear | 157 bp segment of the SARS-CoV-2 spike glycoprotein gene |
| 10 | Hu et al<sup>13</sup> | China | Jinyintan, Hongshan Square Cabin, and Union hospitals in Wuhan | ICU, CT area, staff areas and hallway. | D | WC | AP + O | Not Spec | Ct < 40 (ORF1ab, confirmed with N gene for Ct values between 37 and 40) | ORF1ab, confirmed with N gene |
| 11 | Jan et al<sup>31</sup> | USA | COVID-19 referral center in New Jersey | Patient areas, staff areas and departmental equipment | D | - | - | WHO | Ct < 40 (not clear if both genes) | ORF1ab and E genes |
| 12 | Jin et al<sup>18</sup> | China | Jiangjunshan Hospital in Guizhou | ICU | A | WC | AP + O | WHO | Ct < 40 in either gene = positive and both genes = intensely positive | ORF1ab and NP genes |
| 13 | Kenarkoohi et al<sup>32</sup> | Iran | Shahid Mustafa Khomeini Hospital | ICU and other areas of the hospital | D | Im | AB | - | Ct < 40 (not clear if both genes) | ORF1ab and N genes |
| 14 | Lednický et al<sup>33</sup> | USA | A clinic within a university student health care center | In a hallway, approximately 3 meters from nearest patient traffic. | D | CG | O | - | Not clear | N gene |
| 15 | Lednický et al<sup>14</sup> | USA | University of Florida Health, Shands Hospital, Gainesville, Florida | Designated COVID-19 ward | D | CG | AB | - | Ct < 39.15 | N gene |
| 16 | Li et al<sup>17</sup> | China | Union Hospital, Tongji Medical College, Huazhong University of Science and Technology | ICU ward, general isolation wards, fever clinic and other areas | D | Im | AP + AB | OW | Ct of less than 37 denoted positive findings. A Ct of greater than 37 was subjected to retesting. | Not specified |
| 17 | Liu et al<sup>34</sup> | China | Renmin Hospital of Wuhan University | Field hospital and ICU, plus public areas. | D | GF | AB | - | Ct > 40 (not clear if both genes) | ORF1ab and N genes |
| 18 | Ma et al<sup>20</sup> | China | Two hospitals in Beijing | Covid wards, quarantine hotels, homes | A | WC | AP + AB + O | OW | Ct values of less than 37 or those detected with a Ct value of 37-40 along with an “S” shaped amplification curve. | ORF1ab and N genes |
| 19 | Moore et al<sup>35</sup> | UK | Eight acute hospital trusts in England | Negative pressure isolation rooms | A | WC / GF | AP | WHO | Amplification detected in both replicates. Suspect samples were re-analysed. | ORF1ab, RdRp, E, N genes |
| 20 | Mouchtouri et al<sup>8</sup> | Greece | Various settings | A ferryboat, a nursing home and three COVID-19 isolation hospital wards | D | GF | AB + O | WHO | Not clear | E gene |
| 21 | Ong et al<sup>15</sup> | Singapore | Outbreak center in Singapore | Isolation ward | D | DF / GF | AP + AB + O | OW | Not clear | E gene, although not clear |
| 22 | Razzini et al<sup>36</sup> | Italy | Hospital in Milan | Covid ward | D | GF | AP + AB + O | OW | Ct < 40 | Unclear |
| 23 | Santarpia et al<sup>9</sup> | USA | University of Nebraska Medical Center | Hospital Biocontainment Unit and National Quarantine Unit | D | GF | AP + P | OW | Ct < 39.2 | E gene |
| 24 | Shin et al<sup>37</sup> | Korea | Chungbuk National University Hospital | Mother and daughter in a community treatment centre | A | - | - | OW | Ct < 40 for all three genes | RdRp, N and E genes |
| 25 | Tan et al<sup>38</sup> | China | Tongji Hospital, Huazhong University of Science and Technology | Isolation wards and the intensive care unit (ICU) | D | DF | AP + O | OW | Not clear | ORF1ab |
| 26 | Wang et al<sup>39</sup> | China | First Affiliated Hospital of Zhejiang University | Hospital isolation and ICU | D | - | - | WHO | Ct < 40 | Not specified |
| 27 | Wang et al<sup>40</sup> | China | Wuhan Leishenshan Hospital in Wuhan | ICU and general ward in a field hospital | D | - | - | OW | Not clear | Not specified |
| 28 | Wu et al<sup>41</sup> | China | Wuhan No. 7 Hospital | Hospital isolation and ICU | D | - | - | WHO | Ct < 43 (not clear if both genes) | RdRp and N genes |
| 29 | Ye et al<sup>42</sup> | China | Zhongnan Medical Center of Wuhan University | Various locations | D | - | - | OW | Ct < 40 (not clear if both genes) | ORF1ab and N genes |
| 30 | Zhou et al<sup>43</sup> | UK | A large North West | Various locations | D | WC | U | OW | Duplicate samples | E gene |
|  |  |  | London teaching hospital |  |  |  |  |  | both with Ct < 40.4 |  |
| 31 | Zhou et al<sup>44</sup> | China | Four hospitals in Wuhan, China | Various locations | A | WC | AP + AB + O | OW | Ct < 40 with an "S" shape amplification curve. | ORF1ab and N genes |
| 32 | Declementi et al<sup>12</sup> | Italy | A Trauma Center in Northern Italy | Non-intensive care for Covid patients | D | DF | - | Dry | Not clear | Not specified |
| 33 | Lei et al<sup>45</sup> | China | The First Affiliated Hospital of Guangzhou Medical University | ICU and an isolation ward | A | Im / WC | AP + O | WHO | Not clear | ORF1ab and N genes |
| 34 | Dumont-Leblond et al<sup>46</sup> | Canada | Institut Universitaire de Cardiologie et Pneumologie de Quebec | Acute care hospital rooms | D | DF / GF | AP | - | Either gene Ct <40 | ORF1b and N genes |
| 35 | Moreno et al<sup>7</sup> | Spain | Public transport in Barcelona | Busses and the subway | D | DF | O | Dry | Not clear | NA polymerase (IP2 and IP4) and E gene |

Aim Codes: D = descriptive; A = analytic
Air sampling method: CG = water-based condensational growth; DF = dry filter; GF = gelatin filter; Im = impactor; WC = wetted cyclone
Air sampling strategy: AP = area sample near patient; AB = area sample in the background; O = other location; P = personal; T = in tented enclosure around patient; U = uncertain
Surface sampling codes: WHO = WHO method with wetted swab; OW = other wetted swab; Dry = dry swab; Not Spec – not specified

**Table 2:**
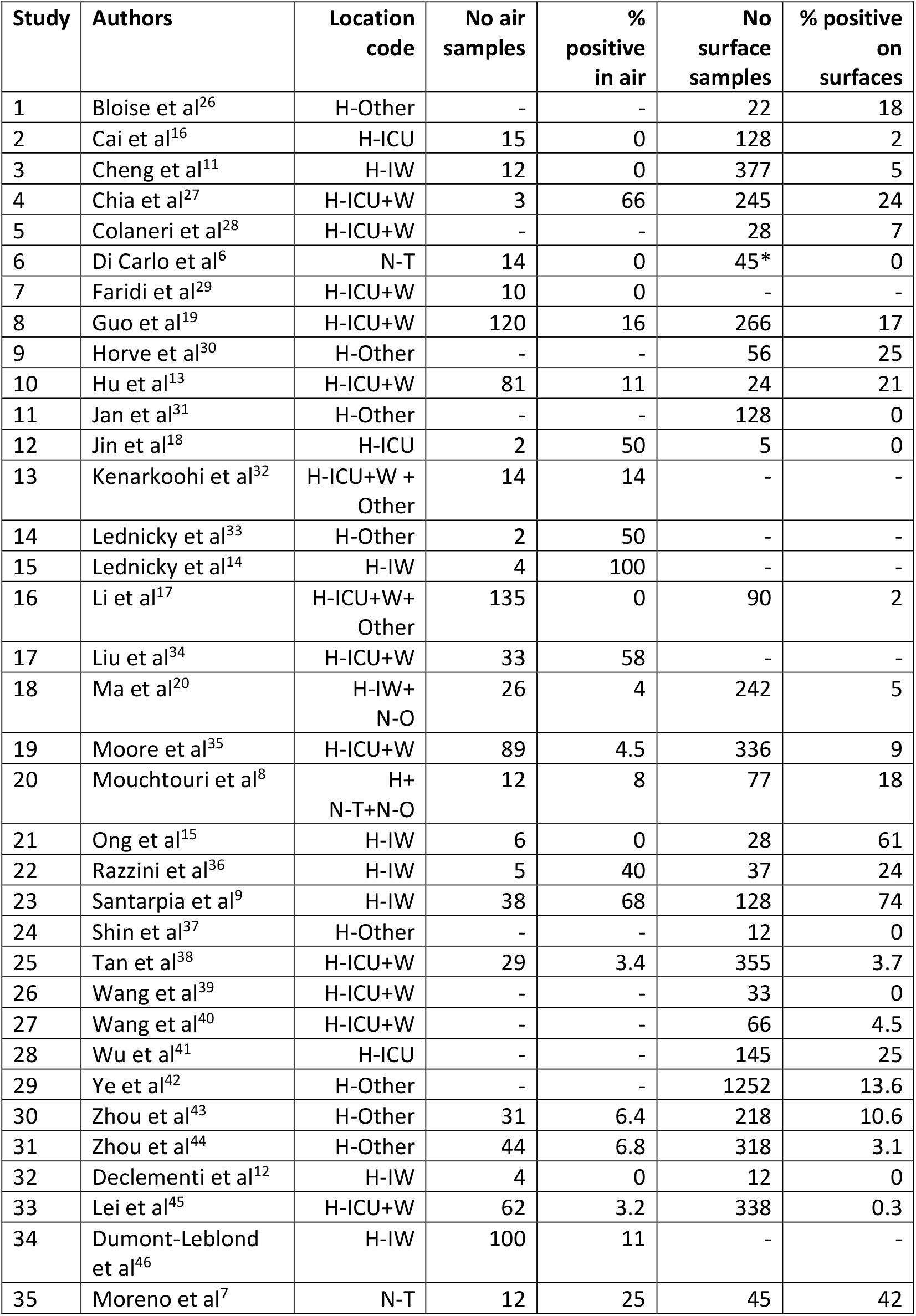
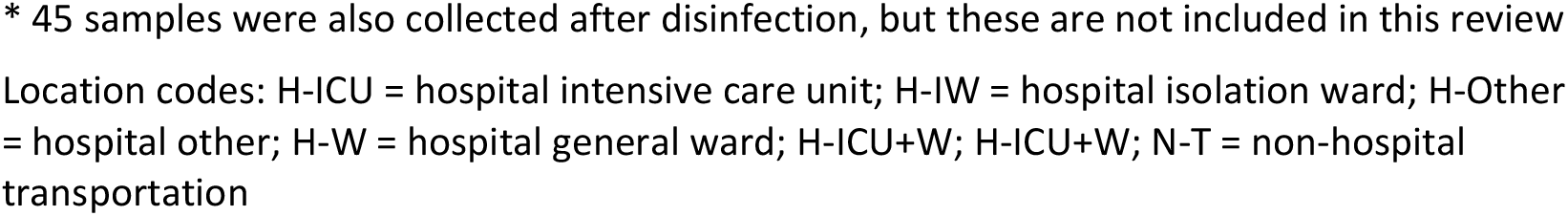
**Air and surface contamination data: number of samples collected and percent positive for SARS-CoV-2 RNA**

| Study | Authors | Location code | No air samples | % positive in air | No surface samples | % positive on surfaces |
| --- | --- | --- | --- | --- | --- | --- |
| 1 | Bloise et al<sup>26</sup> | H-Other | - | - | 22 | 18 |
| 2 | Cai et al<sup>16</sup> | H-ICU | 15 | 0 | 128 | 2 |
| 3 | Cheng et al<sup>11</sup> | H-IW | 12 | 0 | 377 | 5 |
| 4 | Chia et al<sup>27</sup> | H-ICU+W | 3 | 66 | 245 | 24 |
| 5 | Colaneri et al<sup>28</sup> | H-ICU+W | - | - | 28 | 7 |
| 6 | Di Carlo et al<sup>6</sup> | N-T | 14 | 0 | 45* | 0 |
| 7 | Faridi et al<sup>29</sup> | H-ICU+W | 10 | 0 | - | - |
| 8 | Guo et al<sup>19</sup> | H-ICU+W | 120 | 16 | 266 | 17 |
| 9 | Horve et al<sup>30</sup> | H-Other | - | - | 56 | 25 |
| 10 | Hu et al<sup>13</sup> | H-ICU+W | 81 | 11 | 24 | 21 |
| 11 | Jan et al<sup>31</sup> | H-Other | - | - | 128 | 0 |
| 12 | Jin et al<sup>18</sup> | H-ICU | 2 | 50 | 5 | 0 |
| 13 | Kenarkoohi et al<sup>32</sup> | H-ICU+W + Other | 14 | 14 | - | - |
| 14 | Lednický et al<sup>33</sup> | H-Other | 2 | 50 | - | - |
| 15 | Lednický et al<sup>14</sup> | H-IW | 4 | 100 | - | - |
| 16 | Li et al<sup>17</sup> | H-ICU+W+ Other | 135 | 0 | 90 | 2 |
| 17 | Liu et al<sup>34</sup> | H-ICU+W | 33 | 58 | - | - |
| 18 | Ma et al<sup>20</sup> | H-IW+ N-O | 26 | 4 | 242 | 5 |
| 19 | Moore et al<sup>35</sup> | H-ICU+W | 89 | 4.5 | 336 | 9 |
| 20 | Mouchtouri et al<sup>8</sup> | H+ N-T+N-O | 12 | 8 | 77 | 18 |
| 21 | Ong et al<sup>15</sup> | H-IW | 6 | 0 | 28 | 61 |
| 22 | Razzini et al<sup>36</sup> | H-IW | 5 | 40 | 37 | 24 |
| 23 | Santarpia et al<sup>9</sup> | H-IW | 38 | 68 | 128 | 74 |
| 24 | Shin et al<sup>37</sup> | H-Other | - | - | 12 | 0 |
| 25 | Tan et al<sup>38</sup> | H-ICU+W | 29 | 3.4 | 355 | 3.7 |
| 26 | Wang et al<sup>39</sup> | H-ICU+W | - | - | 33 | 0 |
| 27 | Wang et al<sup>40</sup> | H-ICU+W | - | - | 66 | 4.5 |
| 28 | Wu et al<sup>41</sup> | H-ICU | - | - | 145 | 25 |
| 29 | Ye et al<sup>42</sup> | H-Other | - | - | 1252 | 13.6 |
| 30 | Zhou et al<sup>43</sup> | H-Other | 31 | 6.4 | 218 | 10.6 |
| 31 | Zhou et al<sup>44</sup> | H-Other | 44 | 6.8 | 318 | 3.1 |
| 32 | Declementi et al<sup>12</sup> | H-IW | 4 | 0 | 12 | 0 |
| 33 | Lei et al<sup>45</sup> | H-ICU+W | 62 | 3.2 | 338 | 0.3 |
| 34 | Dumont-Leblond et al<sup>46</sup> | H-IW | 100 | 11 | - | - |
| 35 | Moreno et al<sup>7</sup> | N-T | 12 | 25 | 45 | 42 |

\* 45 samples were also collected after disinfection, but these are not included in this review
Location codes: H-ICU = hospital intensive care unit; H-IW = hospital isolation ward; H-Other = hospital other; H-W = hospital general ward; H-ICU+W; H-ICU+W; N-T = non-hospital transportation

Fifteen of the papers were from studies undertaken in China (14 from the mainland and one from Hong Kong), nine from Europe (two from UK, four from Italy, two from Spain and one from Greece), six from North America (five from USA and one from Canada), and five from Asia (two from Singapore, two from Iran and one from Korea). All but three of the studies were carried out in hospitals, mostly in intensive care settings or isolation wards with Covid-19 patients (75% of the healthcare studies). The three non-healthcare papers describe measurements made on public transportation (buses in Northern Italy^6^ and buses and subway trains in Spain^7^) and various workplaces in Greece (a ferryboat and a nursing home – this paper also included data for three COVID-19 isolation hospital wards and a long-term care facility where 30 asymptomatic COVID-19 cases were located^8^). Most of the studies (77%) aimed to describe the contamination present in the setting investigated and the remainder aimed to investigate the extent of contamination in relation to patient viral load or some other patient-related factors.

There are no standardised methods used for quantification of concentrations of SARS-CoV-2 RNA in the air, and as a consequence there were many different methods used. Twenty-five of the studies involved collection of air samples: nine used gelatin filters to collect the sample, eight used wet cyclone samplers, five used impingers, six used dry filters such as PTFE, and two used a water-based condensational growth sampler; some studies used a combination of the techniques (Table 1). Only one study used a personal sampling methodology^9^. The remainder mostly used various combinations of area sampling close to patients (13 studies), in the background near patients (11 studies) or sampling in other areas (12 studies). The volume of air sampled using these methods varied considerably, from 0.09 m^3^ for a midget impinger operated for one hour, to 16 m^3^ for a wet cyclone operating at 400 l/min for 40 minutes. Most samples were collected over a relatively short time, typically less than one hour, and flowrates varied from 1.5 to 400 l/min.

In contrast with the air sampling, there was greater consistency in the surface sampling methodologies used across the studies. There is a method published by the WHO^10^ that recommends samples be collected using a swab with a synthetic tip and a plastic shaft pre-moistened with viral transport medium (VTM). It is recommended that an area of 25 cm^2^ is swabbed, but no recommendations were made concerning the reporting of results as SARS-CoV-2 RNA per cm^2^. Twenty-nine papers contained data from surface sampling: 12 followed the general approach set out by the WHO and 13 used an alternative pre-moistened swab but with, for example, water, saline or phosphate buffer solution in place of VTM. The remaining studies either used dry swabs that were then transported in VTM^11 12,7^ or did not clearly specify the sampling approach used^13^.

In terms of both air and surface sampling, all of the studies used reverse transcription polymerase chain reaction (RT-PCR) analysis to detect the presence of SARS-CoV-2 virus RNA on the collection media (filter, fluid or swab). Ten of the studies attempted to culture positive samples to assess whether the virus was viable, but only one successfully cultured SARS-CoV-2; this study reported high concentrations of SARS-CoV2 RNA on all four air samples collected^14^. A range of gene regions were used in the RT-PCR analysis (mostly combinations of ORF1ab, RdRp, E and N genes; 8 studies used a single gene, 17 used two genes, 3 used three or more genes and 5 did not clearly specify the gene regions used in the RT-PCR analysis). There was a range of criteria used to identify positive samples based on the cycle threshold values (Ct), which is inversely related to the amount of genome material present, with Ct <29 representing abundant target nucleic acid in the sample and Ct of greater than 38 representing minimal RNA present. The criteria used in the studies to identify positive samples ranged from Ct less than 38 to 43 and in cases of multiple gene assays different strategies for identifying positive tests, e.g. replicate analysis of samples where one test was positive and the other negative. None of the studies had an adequate description of quality assurance procedures for both sample collection, e.g. calibration of airflow rates or collection of blank samples in the field, and laboratory analysis, e.g. analysis of blank and spiked samples. Only six of the studies (18%) had any mention of sampling quality assurance procedures and 15 (44%) mentioned some details of analytical quality control.

Twenty-eight studies had contamination data from surfaces (between 5 and 1,252 swab samples) with between zero and 74% positive (median 6%) for SARS-CoV-2 RNA. Twenty-five studies reported air sampling data for between two and 135 samples; the proportion of samples that were positive ranged from zero to 100% with the median across all studies of 6.6% positive samples. These data are summarised in Figure 2. There were six studies that did not detect SARS-CoV-2 RNA on any air samples and five that did not detect SARS-CoV-2 on surfaces; there are no obvious differentiation between these studies and the others reported here. Four studies where less than ten air samples were collected tended to show a high proportion of positive samples (40% - 100%) and it may be that these data are not representative of general conditions in the sampled environments.

**Figure 2:**
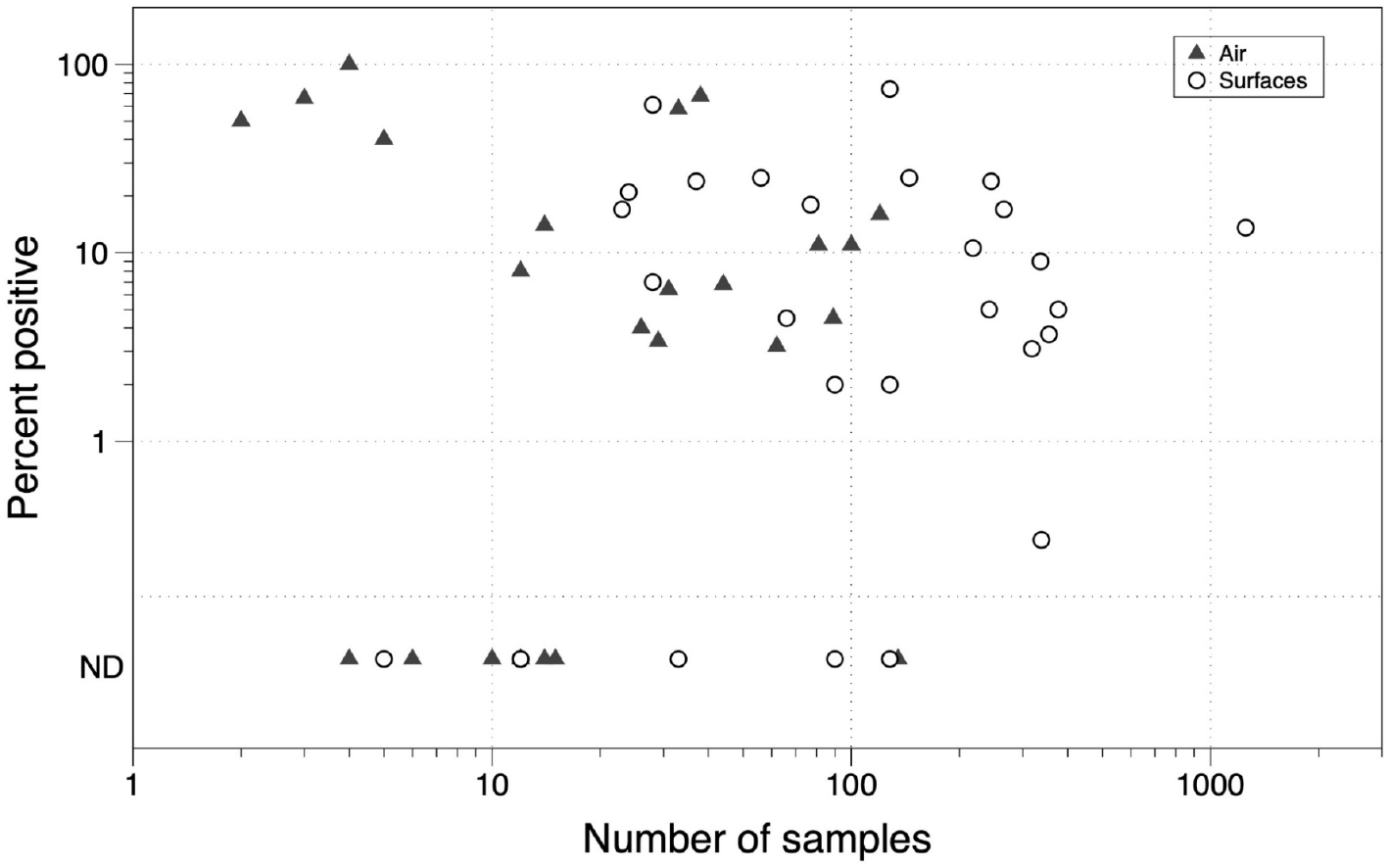
Scatter plot of the percentage of air and surface samples positive for SARS-CoV-2 RNA versus number of samples collected (ND = not detected)

Twenty of the studies had data for both surface and air contamination and these data are summarised in Figure 3, with the area of the data markers proportional to the number of surface samples collected. Note that the double circles represent data from two studies with the same proportion of positive samples. It is clear that in general the proportion of positive samples in a setting was similar for both air and surfaces. There are five outlier studies: four where positive samples were detected on surfaces but not in the air^7,15,11 16 17^ and one study where a relatively high proportion of the air samples were positive but the surface samples were below the limit of detection (LoD)^18^; note this study collected less than 10 samples.

**Figure 3:**
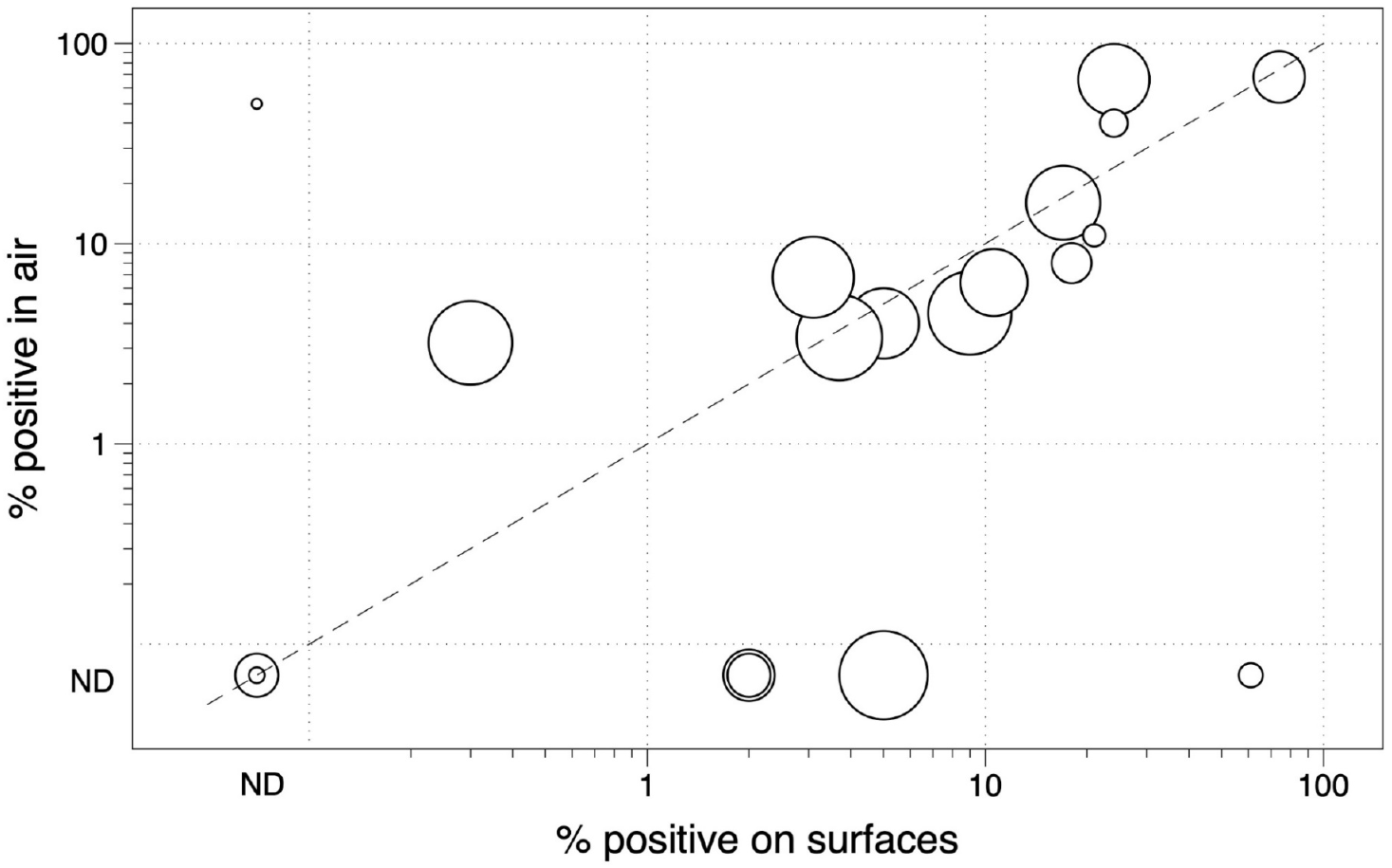
Scatter plot of the proportion of air and surface samples categorised as positive for studies that measured both (ND = not detected)

Only a small number of studies expressed the virus contamination in concentration units. Excluding studies with small numbers of samples (i.e. <10), there were nine that reported air concentrations in terms of virus RNA copies per cubic meter (copies/m^3^). Guo et al^19^ had the largest set of data from an intensive care unit (ICU) and a general Covid ward in Huoshenshan Hospital, Wuhan, China (120 samples obtained between 19 February and 2 March 2020, although the data were only reported as average concentrations for 16 specific locations). They used a wetted wall cyclone that collected air samples at 300 l/min over 30-minute periods. The reported concentrations from three locations in the ICU were between 520 and 3,800 copies/m^3^. However, only 4 of the 26 samples had detectable concentrations and the researchers do not describe how they treated the non-detects when taking the average and is possible that they inappropriately assumed they were zero. At the remaining sampling locations, the results were all below the detection limit, which was unspecified.

Hu et al^13^ provided data from various locations in the Jinyintan, Hongshan Square Cabin, and Union hospitals in Wuhan, collected between the 16^th^ of February to the 14^th^ of March 2020. The concentrations from the positive samples ranged from 1,110 to 11,200 copies/m^3^, although 89% of the measurements were below the LoD. These authors also detected SARS-CoV-2 RNA in some air samples collected outdoors near to the hospital. Data from seven other studies where more than 10 measurements were collected, showed measured air concentrations between around 1 and 8,000 RNA copies/m^3^. We have either used the reported individual data points, extracted data from figures or obtained individual data points through correspondence with the authors, and used these to impute the geometric mean and 95^th^ percentile for each study (Figure 4). Data were available for eight studies: seven from hospital environments and one from transportation (T). In Figure 4, the squares represent the estimated geometric mean, with the size proportional to the total number of air samples. The horizontal line runs between the lower and upper confidence interval for the geometric mean and the vertical line shows the lowest reported measurement, which we assumed as the detection limit. The estimated geometric mean for the whole set of data from hospitals was 0.014 (0.0034-0.047) RNA copies/m^3^.

**Figure 4:**
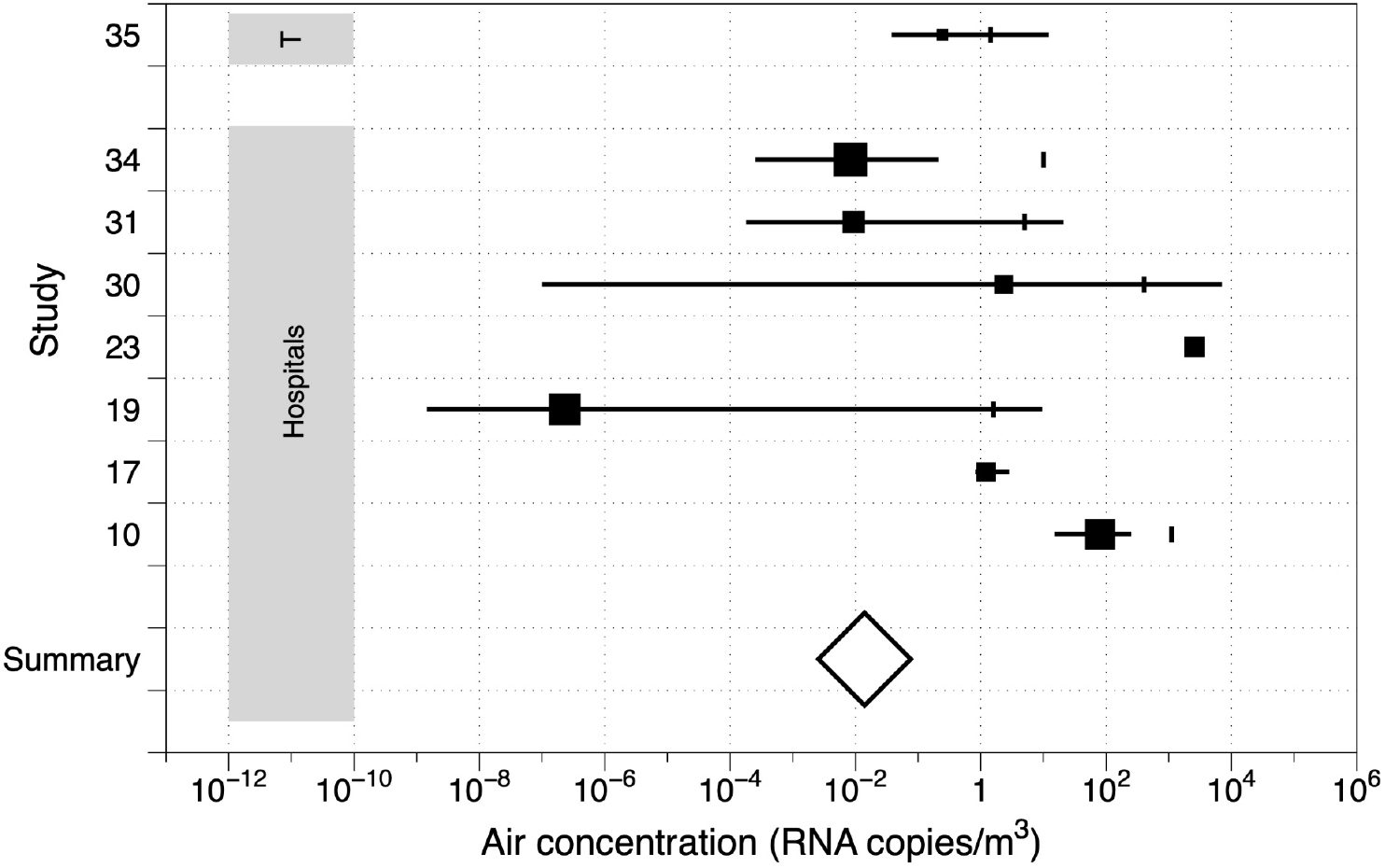
Imputed geometric mean SARS-CoV-2 RNA air concentrations. Note, the black squares represent the imputed geometric mean and the horizontal lines the upper and lower 95% confidence intervals on the geometric mean. The small vertical lines are the lowest reported measurement, which was assumed to be the LoD. T = study in a transportation setting.

Only two studies reported viral loading on surfaces in terms of RNA copies per unit area swabbed^20,7^; where copies were reported otherwise they were expressed per sample collected. Ma and colleagues^20^ collected 242 surface swabs in two hospitals in Beijing and found 4% were positive for SARS-CoV-2 RNA. Loading on the positive samples ranged from 7,100 to 172,000 copies/cm^2^; individual results were not presented. Moreno et al^7^ swabbed surfaces in public transport vehicles in Barcelona. In the subway there were six of 15 swab samples categorised as positive, but for only one of the three target genes analysed. SARS-CoV-2 RNA loading ranged between 0.002 to 0.071 copies/cm^2^ depending on the surface and the target. On the busses 13 of the 30 swabs were positive, mostly for just one gene. Genome loading values ranged between 0.0014 and 0.049 copies/cm^2^.

## DISCUSSION

The studies reviewed here were mostly descriptive and lacked a clear aim other than documenting air or surface contamination. This is perhaps unsurprising given the context of the pandemic and the need to better understand the likely routes of transmission. However, the air sampling methods employed differed greatly between studies, perhaps reflecting local availability of equipment and skills in environmental sampling and previous experience in detecting other airborne viruses, e.g. influenza. Some used methods developed for first SARS outbreak while others used methods adapted for sampling of microbiological exposures, although most air samples were obtained using high volume flowrates over relatively short durations. Almost all of the air samplers had poorly characterised aerosol aspiration efficiencies, i.e. the aerosol size range effectively collected, and cyclone devices likely only effectively sample aerosols with aerodynamic diameter more than around 1μm, e.g. for the WA-400 air sampler Hu et al^13^ quotes a 50% aerodynamic equivalent cut-off diameter of 0.8 μm. However, Liu et al^21^, who collected three samples using a miniature cascade impactor, were able to show the potential for up to half of the SARS-CoV-2 RNA being mainly associated with aerosol with aerodynamic diameter between 0.25 and 1 μm and larger than 2.5 μm. It is therefore possible that many of the studies underestimate the airborne virus RNA concentrations.

In situations where most of the measurements were undetectable, knowledge of the LoD is of prime importance. None of the papers reviewed here provide a clear statement of the LoD in terms of RNA copies per unit of air volume or surface area sampled. Taking the lowest reported value for the air samples, which we accept is only a crude indicator of the LoD, suggests that air concentrations from around 1 to around 2,000 copies/m^3^ were measurable depending on the study. Some, but not all of this variation in the minimum reported air concentrations arises because of variation in the volume of air sampled, which typically ranged from around 0.5 m^3^ to about 16 m^3^. However, there is likely large variation in the sensitivity of the analytical techniques used, e.g. variation in the Ct cut-off value, use of one or more gene sequences for detection and repeat analysis of samples where one gene sequence was undetectable. There is no evidence that the genes used by the different assays would introduce further variation^22^. In clinical testing for SARS-CoV-2 virus in nasopharyngeal swabs, Arnaout et al^23^ noted that the LoD may vary 10,000 fold between approved test kits. Clearly, LoD affects the reporting of positive air and surface samples in the studies reviewed here.

Despite all these limitations, the available data suggests that higher levels of detectable air contamination is associated with higher surface contamination. The most likely explanation for this is that the main source of surface contamination is fine aerosol rather than droplet spray or transfer from the hands of workers or patients. In most healthcare settings the measured airborne concentrations of SARS-CoV-2 virus RNA were low, with likely geometric mean levels around 0.01 RNA copies/m^3^, and the same is undoubtedly the case for surface contamination. The highest concentrations measured in healthcare settings were in excess of 10,000 RNA copies/m^3^ air and around 170,000 RNA copies/cm^2^ surface. Data from public transport settings are limited and there are no data on environmental contamination from other higher risk workplaces such as personal service occupations, factory workers and other non-medical essential workers^24^. Of course, detection of RNA does not mean that there was viable virus present, and in almost all cases the concentration in samples was too low to successfully culture virus. In the one study that successfully cultured virus from four air samples the proportion of virus RNA that was viable ranged from 38% to 79%^14^. It is also important to understand the concentration of viable virus that may give rise to a meaningful level of transmission risk. Karimzadeh et al^3^ estimated that the infective dose of SARS-CoV-19 by aerosol is around 300 virus particles, which if inhaled over a working day might correspond to an average concentration of around 30 SARS-CoV-2 particles/m^3^. There is however a clear need to better understand the infective dose from environmental samples, and whether this differs by the route of intake.

There is a need to develop standardised validated air and surface sampling or analysis methodologies for SARS-CoV-2 RNA, including appropriate quality assurance procedures, to ensure a more comparable set of data across all settings. Researchers should provide detection limits for their analyses in terms of RNA per unit surface area and/or per unit volume of air, and ideally the methods should be developed to reduce the LoD to increase the proportion detectable samples. Where there are data below the LoD, authors should ideally report all of the individual measured values and the imputed geometric mean concentration and other estimated summary statistics and not just the range of detectable concentrations. Ideally, measurements of air concentrations should be representative of long-term personal exposure to inhalable aerosol^25^. Measurement of environmental contamination on its own does not allow a proper interpretation of the exposure of workers, which depends on their interaction with the environment through their personal behaviour. Systematic and uniform reporting of measurement contextual data, e.g. worker behaviour, personal protective equipment worn by worker, room size and ventilation and data on patient status in health and social care setting is crucial to estimation of worker exposure. Similarly, the protocol should specify where and how many samples should be collected in a work environment. International cooperation to establish and maintain such a protocol would facilitate global preparedness for the next outbreak and this task might be appropriately coordinated by the WHO. Understanding environmental transmission early is key to implementation of public health measures to slow the spread of disease throughout work and public/private settings.

## Data Availability

The data extracted from the literature are included in tables in the manuscript.

## ACKNOWLEDGMENTS

We are grateful to a number of our colleagues for comments on early drafts of this paper.

## Footnotes

a Based on 2,769,367 papers listed in WoS for 2020, and 62,478 of these with Covid or SARS-CoV-2 in any data field.

b Based on the papers reviewed here related to the 62,478 papers in WoS with Covid or SARS-CoV-2 in any data field.

